# Examining Cognitive Predictors of Static Balance in Older Adults: The Role of Global Cognition and Inhibition: A cross-sectional study

**DOI:** 10.1101/2025.04.24.25326312

**Authors:** Nahid Divandari, Marie-Louise Bird, Maryam Zoghi, Fefe Vakili, Shapour Jaberzadeh

**Affiliations:** Monash Neuromodulation Research Unit, Department of Physiotherapy, School of Primary and Allied Health Care, Monash University, Australia; School of Health Sciences, University of Tasmania, Australia; Discipline of Physiotherapy, Institute of Health and Wellbeing, Federation University Australia; School of Rural Health, La Trobe University

**Author notes:** **Corresponding author** Nahid Divandari. Funding statement: The authors received no specific funding for this work. Ethical approval and informed consent statements: Ethical approval for this study was obtained from the Monash University Human Research Ethics Committee (MUHREC) and the Human Research Ethics Committee (HREC) at the University of Tasmania. The approval numbers assigned are 31380 for MUHREC and 27343 for HREC at the University of Tasmania. All participants gave their informed consent after receiving thorough information about the study. Participation was voluntary, allowing them the freedom to withdraw at any time. Data availability statement: The data supporting the findings of this study are available from the corresponding author upon reasonable request. Data are available upon request.

**Keywords:** Fall prevention, Mobility, executive function, Physical function, Postural Stability

## Abstract

**Background and Objectives:** Falls are a critical public health issue, with postural balance being key to prevention. While cognition is known to influence balance, the cognitive domain most strongly associated with static balance, and its ability to predict balance performance independently of age, remains unclear.

**Research Design and Methods:** Sixty-two healthy older adults (mean age 74 ± 8.6 years) participated in this cross-sectional study. Static balance was assessed using the Sway Medical App, and cognitive domains were evaluated through global cognition (Mini-Mental State Examination), inhibition (Stroop test), working memory (N-back test), and processing speed (Deary-Liewald test). Hierarchical multiple regression was applied to analyse cognitive predictors of static balance.

**Results:** Higher cognitive performance correlated with better balance. Global cognition and inhibition significantly predicted static balance beyond age, while processing speed and working memory were not significant predictors.

**Discussion and Implications:** Global cognition and inhibition may serve as biomarkers for early detection of balance impairments. Incorporating assessments of global cognition and inhibition into balance evaluations could guide targeted interventions to reduce fall risks and enhance mobility in older adults.

## Introduction

Falls represent an escalating health concern on a global scale, resulting in approximately 37 million hospital admissions annually, with significant costs and lasting impacts on healthcare systems (World Health Organization, 2022). In Australia, falls are the leading cause of hospitalizations due to injuries and also deaths, comprising 43% of all injury-related hospital admissions and 42% of fatalities (Welfare, 2023). Their serious health implications demand immediate, proactive measures that prioritize prevention (Misaghian et al., 2024; Vaishya & Vaish, 2020). Proactively addressing fall risks involves measuring key factors, with balance and cognitive assessments serving as critical components in evaluating fall risk (Chantanachai et al., 2022; Shao et al., 2023; Zhao et al., 2018). This study focuses on the predictability of balance using cognitive assessments to enhance early identification of balance vulnerabilities and inform targeted fall prevention interventions.

Balance is the ability to keep the body’s centre of mass within the base of support (Shumway-Cook & Woollacott, 2007). Static standing balance is a fundamental motor function crucial to the daily life and is strongly associated with a history of falls among older adults (Poncumhak et al., 2023). Its maintenance requires the integration of sensory information from the vestibular, visual, and proprioceptive systems by central nervous system integration (Song et al., 2021). Moreover, cognitive processing which includes the acquisition, analysis, retention, and recall of information, plays a vital role in maintaining balance (Lawlor, 2002; Nazrien et al., 2024). Subtle declines in cognitive function have been linked to balance impairments highlighting the intricate connection between cognition and balance (Chantanachai et al., 2022; Kannan et al., 2022; Muir-Hunter et al., 2014).

Research has examined the connections between cognitive functions—such as executive function, memory, processing speed, and general cognition—and static balance (Divandari et al., 2023; Netz et al., 2018; Redfern et al., 2019). However, there remains a gap in pinpointing which cognitive domains are the most reliable predictors of static balance, especially in older adults. Addressing this gap could enable earlier detection of balance issues and inform targeted interventions that enhance static balance, ultimately supporting fall prevention efforts. Static balance creates a stable base for initiating movement. For example, before taking a step, we naturally stabilize ourselves in a balanced position, which helps prevent falls when transitioning between static and dynamic positions. This stability is particularly critical for older adults, as compromised static balance has been linked to higher fall risks (Poncumhak et al., 2023). Building on evidence that combined cognitive and physical training enhances static balance, we propose that targeted cognitive strategies for improving static balance may foster safer transitions, better balance and help with fall prevention.

This study investigates which cognitive domains are most sensitive in predicting static balance performance beyond the effect of age. By pinpointing the specific cognitive factors that influence balance, this research aims to enhance early identification of balance vulnerabilities and guide targeted interventions to improve stability in older adults.

While gold-standard force platforms and motion capture systems are highly sensitive and reliable for characterizing postural control, their high cost and lack of portability limits clinical use. A newly developed clinically relevant application that is delivered on a phone (SBMA) offers a portable and objective method for measuring postural stability and is used in this study to ensure that the findings can be translated into practice easily by calculating a balance score based on inertial acceleration, which detects postural sway. It measures stability using the built-in motion sensors of any mobile device or tablet to quantify postural sway. Previous studies have demonstrated the reliability of SBMA (Jeremy et al., 2014; Mummareddy et al., 2020).

The primary aims of this study are: 1. to examine the potential association between cognitive domains (executive function, processing speed and general cognition) and static balance, and 2. to identify which cognitive domain significantly predict static balance beyond common confounder of age.

## Methods

### Participants

This is an observational cross-sectional study involving 62 participants, all aged 60 or older, who live independently in the community and can walk unaided. A physiotherapist conducted the data collection over a 90-minute session at a private practice. The exclusion criteria for this study were as follows: 1. presence of any neurological, psychological, orthopaedic, or cardiorespiratory conditions, and 2. experiencing pain that could impact their ability to walk or stand. Ethical approval for this study was obtained from Monash University the Human Research Ethics Committee (MUHREC) and the Human Research Ethics Committee (HREC) at the University of Tasmania. The approval numbers assigned are 31380 for MUHREC and 27343 for HREC. All participants gave their informed written consent after receiving thorough information about the study. Participation was voluntary, allowing them the freedom to withdraw at any time.

### Measurements

Demographic data were collected, including participants’ age, gender, educational background, and history of falls. Body Mass Index (BMI) was measured using bioelectrical impedance with the Seca 804 Flat Scale, which is equipped with chrome electrodes. Cognitive assessments were conducted before the balance tests. Participants began by selecting a random number from 1 to 4 to determine their starting assessment, repeating this process for each subsequent cognitive test. Following the cognitive assessments, participants moved on to the balance tests, which were also administered in a randomized order.

#### Tools for assessment of cognitive domains

***Global cognitive function*** was evaluated with the Mini-Mental State Examination (MMSE) (Folstein, 1975). This test includes 30 questions covering orientation, attention, memory, language, and command-following skills, with scores ranging from 0 to 30 (Nagaratnam et al., 2022).

***Domain-specific cognition*** was assessed using Psytoolkit (Stoet, 2017). This is a free web-based platform for conducting general and psycholinguistic experiments involving complex response time tasks (Kim et al., 2019; Stoet, 2010). The tests were as follows:

***Deary-Liewald reaction time task (DLRT):*** measures reaction time using four white squares on a computer screen, each linked to a specific key (“z,” “x,” “comma,” “full-stop”) (Deary et al., 2011). Participants quickly press the corresponding key when a cross appears in a square, which then disappears, and another cross appears. The inter-stimulus interval varies from 1 to 3 seconds across 8 practice trials and 40 test trials. Median reaction times (ms) are analysed (Figure 1A) (Deary et al., 2011). It demonstrates reliable moderate to excellent test-retest consistency (Ferreira et al., 2021)

**Figure 1.**
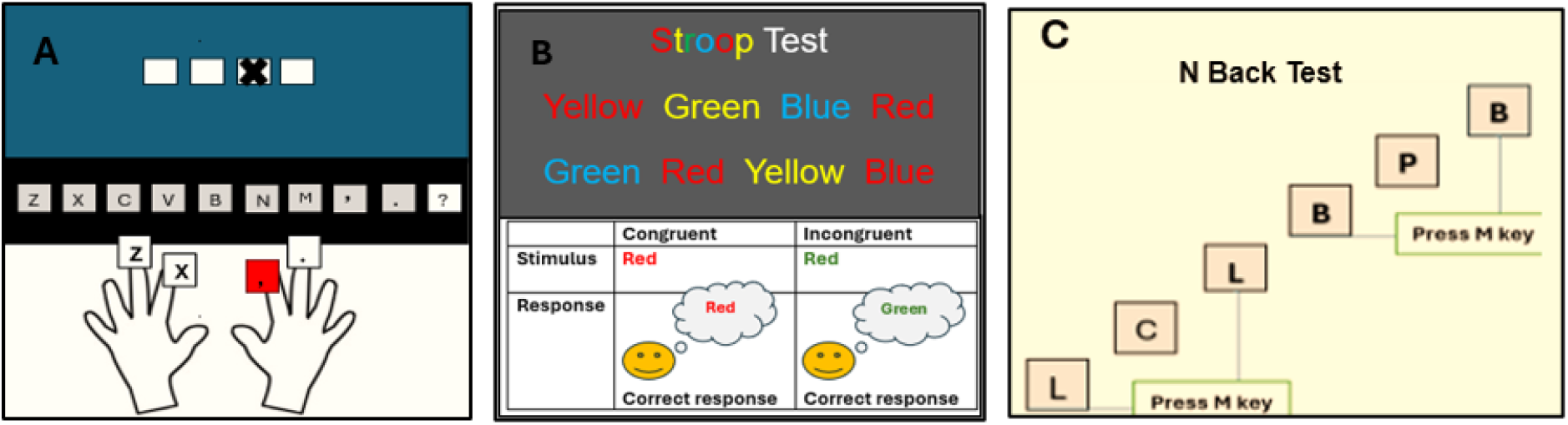
A: Deary-Liewald test B: Stroop test, C:N-back test

***Stroop Color–Word Test:*** measures the ability to inhibit cognitive interference (Scarpina & Tagini, 2017; Stroop, 1935), which are either congruent or incongruent (Figure 1B).Reaction times in incongruent conditions are analysed for interference effects (Periáñez et al., 2020).

***N-Back Test:*** assesses working memory (Kirchner, 1958). Participants view a series of stimuli and identify if the current stimulus matches the one from three trials earlier (Frost et al., 2021). It includes 100 trials, and the accuracy of responses is analysed (Figure 1C) (Gajewski et al., 2018).

##### Balance tests

The static balance test was conducted using the Sway Balance Mobile Application (SWAY; Sway Medical, Tulsa, OK) (Vincenzo et al., 2016). This FDA-approved, reliable, and validated method assesses postural stability through accelerometers (Jeremy et al., 2014). Participants held the device over their sternum, and the accelerometers recorded movement during single-leg and tandem stances, both with eyes open and closed.

The app converted the data into balance scores ranging from 0 to 100, with lower scores reflecting poorer balance (Amick et al., 2013). Each test comprised a practice trial and three repetitions, with the average score calculated. Positions were timed for 10 seconds with a 3-second countdown. (Vincenzo et al., 2016). If participants got out of the position and touched the floor, the trial scored zero.

#### Data management and analysis

Descriptive analysis of all participants demographic data. Then, the distribution of all variables was checked for normality using Shapiro-Wilk Test (p<0.05). Pearson’s correlation coefficients for all variables were computed to evaluate the interrelationships between them (Gatto et al., 2020) (Table 1). Among potential confounders, age was selected for inclusion in the regression models, as it exhibited significant correlations with most cognitive and balance variables, unlike other variables such as gender, education, falls history, BMI, and body fat mass, which were not found to be significant confounders. These variables were excluded as they did not show significant correlations with the primary variables of interest and did not meaningfully contribute to the variance in balance outcomes. To investigate whether cognition contributes to a significant portion of the variance in static balance beyond age, hierarchical multiple regression analysis (MRA) was conducted (p<0.05, 0.01, & 0.001) (Tabachnick, 2019). MRA was selected because it allows for the assessment of how much additional variance in balance performance is explained by cognitive domains after accounting for age. This method also facilitates the stepwise inclusion of variables to investigate their unique contributions. In this case, age was entered first into the model as a predictor, as it had strong correlations with balance measures. Each cognitive domain was then added in subsequent steps to evaluate whether it significantly increased the explained variance (measured as a significant change in R^2^) in relation to the balance tests. The analysis involved four models, repeating the process for each cognitive domain in relation to the balance tests. In the first step, age was evaluated as predictors. In the second step, each cognitive domain was added individually to assess whether it significantly increased the explained variance (noted as a significant change in R^2^) of the model (Tabachnick, 2019). Prior to interpreting the MRA results, several assumptions were assessed. Stem-and-leaf plots and boxplots were used to verify the normal distribution and absence of univariate outliers in each regression variable. Normal probability plots of standardized residuals and scatterplots of standardized residuals against standardized predicted values were examined to confirm the assumptions of normality, linearity, and homoscedasticity of residuals were met (Tabachnick, 2019).The sample size was determined using G power (Faul et al., 2007).

**Table 1.**
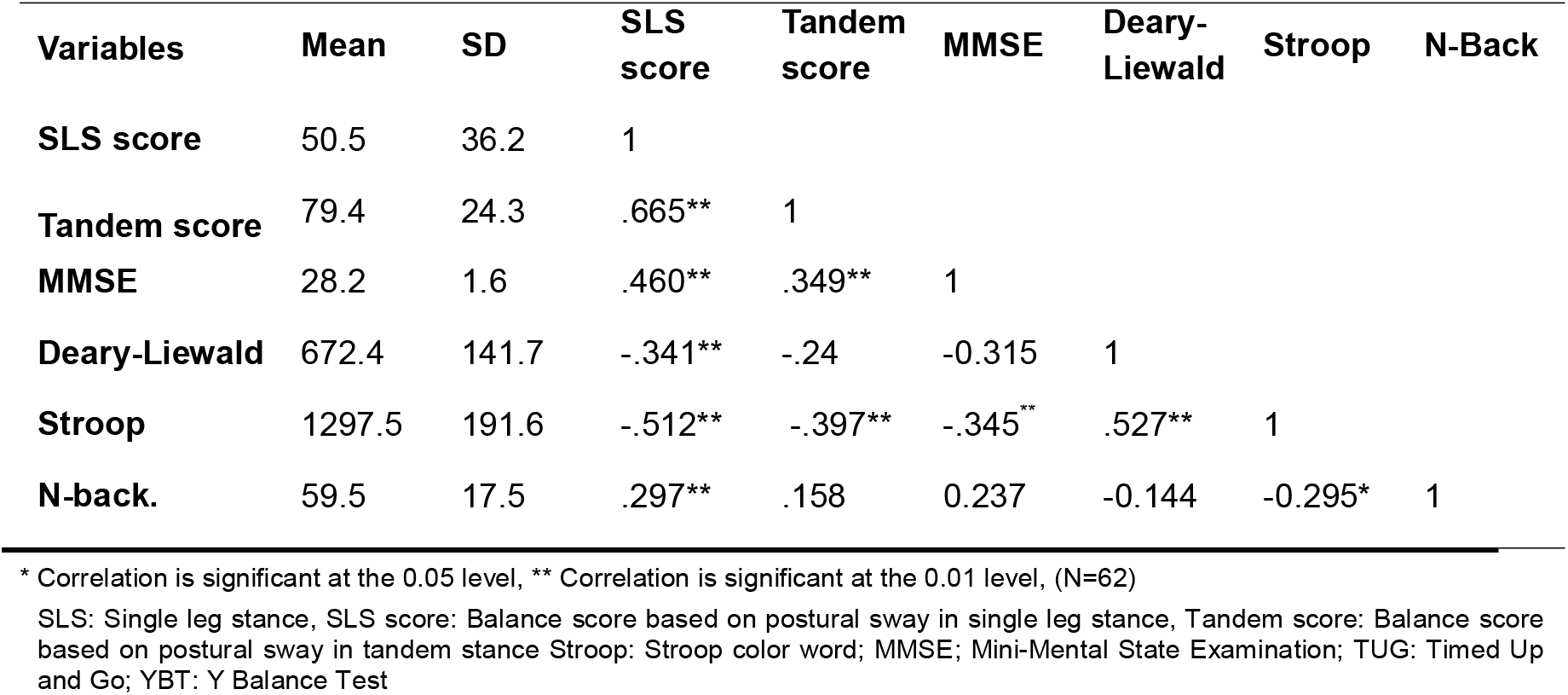
Descriptive statistics for cognitive and balance measures (N=62) and correlations between cognitive domains and balance.

## Result

### The descriptive analysis

A total of 62 older adults participated in this study. Participants ranged in age from 65 to 85 years, with a mean body mass index (BMI) of 28 (SD = 5.7). Approximately two-thirds of the sample were female. Just over half had completed tertiary education. Additionally, 40% of participants reported at least one fall in the past 12 months.

Baseline cognitive and balance test results are detailed in Table 2. Global cognition results showed skewness and were subsequently log-transformed.

**Table 2.** Hierarchical regression results for cognitive domains predicting static balance

| Model 1 |  |  | Model 2 |  |  |
| --- | --- | --- | --- | --- | --- |
| Independent Variables | SLS | Tandem | Independent Variables | SLS | Tandem |
| Step 1: Background variables |  |  | Step 1: Background variables |  |  |
| Age | -.565*** | -.435*** | Age | -.565*** | -.435*** |
| Step 2: Cognitive variable |  |  | Step 2: Cognitive variable |  |  |
| Age | -.404** | .291* | Age | -.463** | -.439* |
| Inhibition | .286* | .258 | Global cognition | .298** | .252* |
| R<sup>2</sup> step 1 (Age) | .322 | .191 | R<sup>2</sup> step 1 (Age) | .322 | .191 |
| R<sup>2</sup> step 2 (Age+ Inhibition) | .372 | .231 | R<sup>2</sup> step 2 (Age+ global cognition) | .399 | .246 |
| R<sup>2</sup> change | .05* | .04 | R<sup>2</sup> change | .077** | .055** |

| Model 3 |  |  | Model 4 |  |  |
| --- | --- | --- | --- | --- | --- |
| Independent Variables | SLS | Tandem | Independent Variables | SLS | Tandem |
| Step 1: Background variables |  |  | Step 1: Background variables |  |  |
| Age | -.565*** | -.435*** | Age | -.565*** | -.435*** |
| Step 2: Cognitive variable |  |  | Step 2: Cognitive variable |  |  |
| Age | -.530*** | -.419*** | Age | -.535*** | -.420*** |
| Working memory | .193 | .288 | Processing speed | -.060 | -.030 |
| R<sup>2</sup> step 1 (Age) | .322 | .191 | R<sup>2</sup> step 1 (Age) | .322 | .191 |
| R<sup>2</sup> step 2 (Age+ Working Memory) | .355 | .198 | R<sup>2</sup> step 2 (Age+ Processing speed) | .325 | .191 |
| R<sup>2</sup> change | .033 | .007 | R<sup>2</sup> change | .002 | .001 |

Age and cognitive domains are predictors. SLS: static balance score based on postural sway in single Leg Stance position, Tandem: static balance score based on postural sway in tandem position; $\beta$ : standardized regression coefficient; $R^2$ : The coefficient of determination; $R^2$ change improvement in R-square when the second predictor is added, \* $P < 0.05$ \*\* $P < 0.01$

### Association of demographic information with cognitive and balance measures

Age was significantly associated with all static and balance tests data and cognitive measures, except for working memory. Other demographic variables did not show significant correlations with cognitive and balance measures.

### Association between cognitive domains and balance measures

All cognitive domains, including global cognition, inhibition, working memory, and processing speed, exhibited significant moderate associations with balance scores based on postural sway during single-leg stance (correlation coefficients ranging from -0.512 to 0.497, p < 0.01). Both global cognition and inhibition showed significant moderate association (ranging from -0.397 to 0.349, p<0.01) with balance score based on postural sway in tandem position (Table 1). However, processing speed and working memory did not show any association with this balance test.

### Cognitive domains predicting static balance beyond the effect of age

The simple associations between cognitive domains and balance measurements shown in Table 1 are not sufficient evidence for functional relationships (Rabbitt et al., 2006). To prove functional causality, it is necessary to test how well the associations between cognitive test scores and balance test scores hold up when accounting for the effects of age, which are significantly correlated with both sets of scores. This was achieved using hierarchical multiple regression analyses (MRA). In the first step, the R^2^ values for predicting balance scores from age were calculated. In the second step, new R^2^ values were obtained after adding one cognitive score to the model. The significance of changes in R^2^ was evaluated to determine if the inclusion of cognitive scores accounted for additional variance in balance test scores. This method was applied to each of the balance tests: balance score based on postural sway in single leg stance, and tandem positions, allowing for a detailed analysis. In the first step, age emerged as significant predictors of balance performance across all tests. Upon adding cognitive domains in the second step, the results were as follows in table 2:

Global cognition, measured by the MMSE, significantly predicted scores on all balance tests (Table 2, Model 2). Inhibition, assessed by the Stroop test, significantly predicted scores on balance scores based on postural sway in the single leg stance position, but not on tandem position sway scores (Table 2, Model 1). Processing speed and working memory did not significantly predict any balance test scores (Table 2, Model 3 and 4 respectively).

### Cognitive domains predict static balance

In this study, hierarchical MRA was conducted with four models, each including age in step one, and adding a cognitive domain in step two, to predict static balance (Table 2). The results are described for both static balance tests in each model as follows:

#### Step 1: background variables

Age significantly predicts balance performance in single leg stance position (F(1,62) = 14.019, P < 0.001, R^2^ = 0.322), and in tandem position (F(1,62) = 6.950, P = 0.002, R^2^ = 0.191), in step one in all four models. The second step in each model for every cognitive domain is as follows:

##### Model 1: Inhibition

For SLS, the overall fit of model 1 was significant at step 1 and remained significant after adding the inhibition score in step 2 (F (2,62) = 11.475, P < 0.001, R^2^ = 0.372). There was a significant change to model fit (ΔR^2^ = 0.050, P < 0.05). A significant change in R2 is illustrated in Figure 3A when inhibition is added to age as a cognitive predictor. For Tandem, the overall fit of model 1 was significant at step 1 and remained significant after adding the inhibition score in step 2 (F (2,62) = 5.803, P = 0.002, R^2^ = 0.231). However, there was no significant change to model fit (ΔR^2^ = 0.040, P >0.05).

##### Model 2: Global cognition

For SLS, the overall fit of model 1 was significant at step 1 and remained significant after adding the global cognition score in step 2 (F(2,62) = 12.825, P < 0.001, R^2^ = 0.399). That accounted for a significant change (ΔR^2^ = 0.077, P = 0.009), which is depicted in figure 3B. When predicting tandem, the overall fit of model 2 was significant at step 1 and remained significant at step 2 (F (2,62) = 6.297, P < 0.001, R^2^ = 0.246). Adding global cognition scores significantly improved the overall model fit (ΔR^2^ = 0.055, P < 0.01). Figure 3C illustrates the incremental change in R^2^ when adding the cognitive predictor of global cognition to age in this MRA model.

##### Model 3: Working Memory

For SLS, the overall fit of model 1 was significant at step 1 and remained significant after adding the working memory scores (N-Back test) scores at step 2 (F (2,62) = 10.663, P < 0.001, R^2^ = 0.355). The addition of working memory scores did not result in a significant change in the overall model fit (ΔR^2^ = 0.033, P > 0.05). When predicting tandem, the overall fit of model 2 was significant at step 1 and remained significant at step 2 (F (2,62) = 4.764, P = 0.005, R^2^ = 0.198). The addition of working memory scores did not result in a significant change in the overall model fit (ΔR^2^ = 0.007, P > 0.05).

##### Model 4: Processing speed

For SLS, the overall fit of model 2 was significant at step one and remained significant after adding processing speed scores (Deary-Liewald) at step 2 (F (2,62) = 9.291, P < 0.001, R^2^ = 0.325). However, adding processing speed did not cause any significant change in overall model fit (ΔR^2^ = 0.002, P > 0.05). Similarly for tandem, the overall fit of model 2 was significant at step 1 and remained significant at step 2 (F (2,62) = 4.573, P = 0.006, R^2^ = 0.191). The addition of processing speed scores did not significantly alter the overall model fit (ΔR^2^ = 0.001, P > 0.05).

## Discussion

In this study, we explored the relationship between various cognitive domains and static balance, aiming to identify the cognitive domain that most significantly influences balance performance, beyond the confounding effects of age. By employing a hierarchical MRA, we were able to isolate the contribution of each cognitive domain to balance, providing insights into whether cognitive factors independently affect balance performance in older adults.

The study’s results confirmed significant positive associations between cognitive function and balance performance. Specifically, individuals with higher cognitive function exhibited better performance on static balance tests. These results align with the current body of literature concerning both healthy and health-compromised older adults (Biasin et al., 2023; Redfern et al., 2019). The MMSE test (evaluating global cognition) and the Stroop test (assessing inhibition) exhibited significant associations with both static balance tests. These are aligned with previous studies (Goto et al., 2018; Muir-Hunter et al., 2014; Rosano et al., 2005). Furthermore, the Deary-Liewald task and N-Back test, which measures processing speed and working memory respectively, demonstrated significant associations with balance score based on postural sway in single leg stance position but not in tandem position (probably because of the ceiling effect in this test). Given the robustness of our population and the test results, it appears that balance test in tandem position was not as challenging as balance test in SLS position.

Hierarchical MRA shows that cognitive function and age together significantly predict static balance tests in older adults. Initially, using age as the predictor demonstrates strong predictive power for balance test scores. However, although all cognitive domains together with age significantly predict balance scores, certain cognitive facets notably increase the R^2^ value, emphasizing their specific independent predictive power. This highlights the critical role of both age and specific cognitive domains in predicting static balance in older adults.

Global cognition has emerged as the strongest predictor of static balance tests among cognitive domains after controlling for age. This was evidenced by a statistically significant increase in the R^2^ value when MMSE scores were added as predictors alongside age. Meta-analyses in 2016 and 2023 found an association between MMSE scores and balance, with the 2023 study specifically linking MMSE to static balance (Demnitz et al., 2016; Divandari et al., 2023). Additionally, Naito et al. (2023) identified a significant association between MMSE scores and postural sway in a single-leg stance position, although in this study, MMSE scores were treated as the dependent variable in the regression analysis (Naito et al., 2023). Khan et al. (2023) reported a significant moderate association between the Berg Balance Scale and MMSE (Khan et al., 2023). Other studies have also revealed a correlation between global cognition and static balance (Goto et al., 2018; Rosano et al., 2005; Tsutsumimoto et al., 2013Won, et al, 2014 reported that within the range of cognitive performances studied, the evaluation of global cognitive performance through MMSE exhibited the most substantial impact from physical function. However, in this study balance was predicting cognition.

Our results differ from those of Zhao et al. (2022), who reported no significant association between one-leg balance with eyes open and MMSE scores. This inconsistency could be due to different population and testing methods, Zhao et al. (2022) recruited participants through community health service centres, whereas our study participants were a robust group. These methodological distinctions highlight the need to consider diverse factors that may contribute to variations in study outcomes and may affect the generalisability of the results.

In terms of the mechanism of MMSE predictability for balance, it is conceivable that age-related alterations in cognition may serve as the driving force behind changes in balance function. A longitudinal study of older adults found that a decline in cognitive function occurred before the onset of mobility limitations (Elovainio et al., 2009). Researchers have investigated that the primary direction of the association is from impaired cognition to diminished physical activity (Gothe et al., 2014).

Furthermore, doing Stepwise regression showed that Stroop test, assessing inhibition, was able to predict balance score in SLS and tandem stance positions even after controlling for age with better score on Stroop test better performance in these balance tests. It showed a statistically significant change in R^2^ after adding Stroop test results to age. This is aligned with previous studies indicating a positive link between better balance and Stroop test performance (Berryman et al., 2013; Zettel-Watson et al., 2017). However, in these studies balance was predicting cognition. Executive function is a predictor mobility outcome (Gothe et al., 2014). Inhibitory control plays a crucial role in balance performance (Kwag & Zijlstra, 2022). Inhibition, a core component of executive function (Boucard et al., 2012) plays a pivotal role in an individual’s capacity to stand, walk, and interact with the environment safely and efficiently (Levin et al., 2014). It is also vital for the sensory integration processes necessary to maintain balance, especially in older adults (Redfern et al., 2009). Age-related decline in postural control is linked to structural changes in the middle frontal gyrus and basal ganglia, brain regions that are crucial for response inhibition across various sensorimotor tasks (Boisgontier et al., 2017). This observation aligns with the understanding that the cognitive inputs crucial for postural control adapt according to the task’s complexity and an individual’s proficiency in postural control abilities, as suggested by previous research (Horak & Macpherson, 2011).

Including processing speed or working memory in the regression models alongside age did not significantly enhance the prediction of balance (no substantial increase in R^2^). Our findings on processing speed are consistent with previous research (Rosano et al., 2005; Won et al., 2014); However, they are inconsistent with Demnitz et al. (2017). This discrepancy may be due to the use of different tests for balance and processing speed in their study. The results of this study on working memory differ from previous research by Kawagoe et al. (2014, 2015) and Mato et al. (2020). Unlike their studies, which utilized face and location working memory tests, we focused on letter working memory. Notably, letter working memory tends to decline less with age compared to memory for faces and objects in healthy older adults (Leonards et al., 2002). This suggests that the decline in visual working memory is not solely due to the increased vulnerability of the prefrontal cortex, which is responsible for executive processing (Leonards et al., 2002). Therefore, it is recommended to avoid using visual (letter) working memory as an outcome measure or treatment component in dual-task tests or rehabilitation exercises. However, further research is essential to differentiate between various types of working memory when investigating their relationship with balance.

Our study underscores the clinical significance of cognitive assessments, specifically the MMSE and Stroop test, in predicting balance impairments among older adults. By identifying cognitive domains such as global cognition and inhibitory control as predictors of balance performance, clinicians can enhance fall-risk assessment and implement early, targeted interventions. This approach supports personalized strategies to mitigate fall risk and improve safety in older This study also underscores the clinical significance of incorporating cognitively demanding balance assessments and interventions to effectively address balance issues in older adults. The significant predictive power of global cognition and inhibition suggests that targeted cognitive training—focusing on general cognitive function and inhibitory control—could enhance the effectiveness of balance interventions. For instance, incorporating dual-task exercises that engage global cognitive function and inhibitory control into standard balance training may enhance both balance and cognitive health, thereby supporting fall prevention and promoting mental resilience in older adults. Furthermore, our findings indicate that different balance tasks engage distinct cognitive resources; specifically, inhibition predicts performance in single-leg stance (SLS) but not in tandem stance, whereas global cognition supports both. This suggests that SLS may benefit from exercises emphasizing attentional control and inhibitory processes, while tandem stance may be less cognitively demanding. These insights highlight the need for further research to refine task-specific cognitive demands, enabling clinicians to design more effective, individualized interventions that align with each patient’s unique needs. Such tailored approaches have the potential to significantly enhance fall prevention strategies, fostering greater functional independence and quality of life in older adults. Also, it is advised to analyse the balance tasks more to understand which aspect of cognition is more involved on task for example standing on SLS or tandem may not require that much processing speed but some other tasks that of balance may require more. Further studies should focus on analysing different types of balance tasks and identifying the specific cognitive resources each requires. This deeper understanding would enable the development of more tailored and effective exercise plans and interventions, ultimately improving outcomes in balance training and fall prevention.

### Limitations

The findings in this study should be interpreted considering the following limitations. Firstly, while we examined the correlation of sex, age, education, and other potential confounders such as falls history, BMI, and body fat mass on the cognitive and balance functions of older adults, there are other factors that may affect cognition and balance that were not accounted for foot pathologies such as flat foot (Febriyanti et al., 2024) decreased foot arch, arthritic changes in lower limbs (John et al., 2024). Secondly, this study focused on a population of healthy older adults, which may limit the generalizability of the results to older adults with health conditions or impairments. Including older adults with varied health profiles in future studies could offer insights into how cognitive and physical declines interact in more diverse populations.

## Conclusions

In summary, this study explored the interplay between cognitive domains and static postural balance in older adult population. Global cognition and inhibitory control emerged as significant predictors of static balance in older adults, independent of age, emphasizing the critical role these cognitive domains play in maintaining postural stability. The results suggest the potential suitability of postural sway score in SLS position for assessing the cognitive-balance relationship in robust older populations over tandem test. Also, we propose the integration of MMSE and Stroop tests into routine balance assessments and treatments by physiotherapists. The integration of cognitive assessments into balance evaluations, particularly focusing on global cognition and inhibitory control, could enhance the precision of fall risk assessments in older adults. By incorporating tools like the MMSE and Stroop test, clinicians may detect cognitive impairments that contribute to balance issues earlier, allowing for more personalized intervention strategies. These findings suggest that a more holistic approach to rehabilitation, one that addresses both cognitive and physical components, could reduce fall risks and improve functional independence in older adults.

## Data Availability

All data produced in the present study are available upon reasonable request to the authors

## Acknowledgement

We would like to acknowledge Dr. Tim Power, Consultant in Data Science, AI, and Sensitive Data Platforms at the Monash Research Centre, for his invaluable assistance with the statistical analysis in this paper. We also extend our gratitude to Miss Fatemeh Vakili for her exceptional help in data collection process.

## Notes

Declaration of conflicting interest: All authors have contributed significantly to the development of this manuscript and have reviewed and approved the final version of the manuscript, taking full responsibility for its content. The authors confirm that there are no financial, personal, or professional affiliations or relationships that could be perceived as influencing the research presented in this manuscript.

### Competing Interest Statement

The authors have declared no competing interest.

### Funding Statement

This study did not receive any funding

### Author Declarations

Ethical approval for this study was obtained from Monash University the Human Research Ethics Committee (MUHREC) and the Human Research Ethics Committee (HREC) at the University of Tasmania. The approval numbers assigned are 31380 for MUHREC and 27343 for HREC. All participants gave their informed written consent after receiving thorough information about the study. Participation was voluntary, allowing them the freedom to withdraw at any time.

